# Early Impact of COVID-19 Pandemic on Paediatric Surgical Practice in Nigeria: a National Survey of Paediatric Surgeons

**DOI:** 10.1101/2020.05.24.20112326

**Authors:** I.O. Ogundele, F.M. Alakaloko, C.C. Nwokoro, E.A. Ameh

**Author notes:** **Corresponding Author:** Ibukunolu O. Ogundele, Department of Surgery, Olabisi Onabanjo University Teaching Hospital, PMB 2001, Sagamu, Ogun State, Nigeria. E-mail address. **Competing Interests**: All authors declare no conflicts of interest. **Funding**: This research received no specific grant from any funding agency in the public, commercial or not-for-profit sectors. **Contributorship**: IOO and FMA conceived the research idea. IOO, FMA and CCN conducted the survey. IOO, FMA and EAA made the drafts of the manuscript. All the authors made substantive contributions to the intellectual content and reporting of the work described in this article. They all read and approved the final manuscript. IOO and FMA are responsible for the overall content as guarantors. **Ethics Approval**: Health Research Ethics Committee of Olabisi Onabanjo University Teaching Hospital, Sagamu, Ogun State, Nigeria. OOUTH/HREC/339/2020AP.

## Abstract

**Introduction:** The novel Coronavirus disease has had significant impact on healthcare globally. Knowledge of this virus is evolving, definitive care is not yet known, and mortality is increasing. We assessed its initial impact on paediatric surgical practice in Nigeria, creating a benchmark for recommendations and future reference.

**Methods:** Survey of 120 paediatric surgeons from 50 centres to assess socio-demographics and specific domains of impact of COVID-19 on their services and training in Nigeria. Seventy four surgeons adequately responded. Responses have been analysed. Duplicate submissions for centres were excluded by combining and averaging the responses from centres with multiple respondents.

**Results:** Forty-six (92%) centres had suspended elective surgeries. All centres continued emergency surgeries but volume reduced in March by 31%. Eleven (22%) centres reported 13 suspended elective cases presenting as emergencies in March, accounting for 3% of total emergency surgeries. Nine (18%) centres adopted new modalities for managing selected surgical conditions: non-operative reduction of intussusception in 1(2%), antibiotic management of uncomplicated acute appendicitis in 5(10%), more conservative management of trauma and replacement of laparoscopic appendectomy with open surgery in 3(6%) respectively. Low perception of adequacy of Personal Protective Equipment (PPE) was reported in 35(70%) centres. Forty (80%) centres did not offer telemedicine for patients follow up. Twenty-nine (58%) centres had suspended academic training. Perception of safety to operate was low in 37(50%) respondents, indifferent in 24% and high in 26%.

**Conclusion:** Majority of paediatric surgical centres reported cessation of elective surgeries whilst continuing emergencies. There is however an acute decline in the volume of emergency surgeries. Adequate PPE need to be provided and preparations towards handling backlog of elective surgeries once the pandemic recedes. Further study is planned to more conclusively understand the full impact of this pandemic on children’s surgery.

**What is already known and what this study adds:** Anecdotal evidence suggests that elective surgeries in children have been suspended due to COVID-19 pandemic.
Our study shows that most centres have suspended elective surgeries. All centres continued emergency surgeries but the volume reduced by 31% in March 2020. Moreover, 3% of the emergency surgeries were some of the suspended elective cases presenting as emergencies. Almost 20% of centres have newly adopted non-operative modalities for managing selected emergency surgical conditions.
This data shows an urgent need for consensus guidelines for emergency services and protocols for handling backlog of elective surgeries in children once the pandemic recedes. Outcome of the modifications in treatment may be subject to future research.

## BACKGROUND

Corona virus disease 19 (COVID-19) is a highly transmissible novel viral illness, caused by severe acute respiratory syndrome coronavirus 2 (SARS-CoV-2) (1). It was reported to have emerged in Wuhan, China, in December 2019 but later spread to other parts of China and other countries of the world (2). This disease poses a huge challenge to health care systems around the world. The U.S. Department of Health and Human Services stated in its 2017 Pandemic Influenza plan update that “emerging viral pandemics can place extraordinary and sustained demands on public health and health systems and on providers of essential community services” (3). The effect may be more profound in regions with already limited resources and fragile health infrastructure. The aim of this study was to carry out a survey of paediatric surgeons in a resource limited setting to assess early effects of the COVID-19 pandemic on their practice in the initial stages of the outbreak. Data obtained would be used for recommendations and future reference.

## METHODS

Relevant information was obtained from paediatric surgeons (consultants and senior registrars) currently practising in Nigeria, using a pre-tested questionnaire designed on Microsoft Word version 10 (Microsoft Seattle, WA, USA) and transcribed to google form. The questions were based on 5-point Likert scale (Strongly agree, agree, neither agree nor disagree, disagree, strongly disagree). We circulated the forms to the predetermined group of specialists by email and online chat rooms and kept them open from 10th to 17th April 2020. Daily reminders were also sent.

Participants were required to provide socio-demographic data, information on patient traffic and decision on management of specific conditions, availability of PPE, impact on surgeon’s psyche, their academic programs and institutions infrastructure.

A total of 120 paediatric surgeons were sent the survey. Eighty-three paediatric surgeons responded but 74 were adequately completed. For the purpose of analysis, the 5-point Likert scale was reduced to 3 points (Agree, neutral, disagree).

Duplicate submissions for centres were excluded by combining and averaging the responses from centres with multiple responses.

Responses were analysed using SPSS version 22 and presented as categorical data and percentages.

### Patient and Public Involvement statement

This research was done without patient involvement. Patients were not invited to comment on the study design and were not consulted to develop patient relevant outcomes or interpret the results. Patients were not invited to contribute to the writing or editing of this document for readability or accuracy.

### Ethics Approval

Obtained from the Health Research Ethics Committee of Olabisi Onabanjo University Teaching Hospital, Sagamu, Ogun State, Nigeria (00UTH/HREC/339/2020AP).

## RESULTS

### Demographics

The response rate was 74(61%). The 74 completed responses represented 50 centres across the country. Table 1 shows the socio-demographic characteristics of respondents.

**Table: 1.**
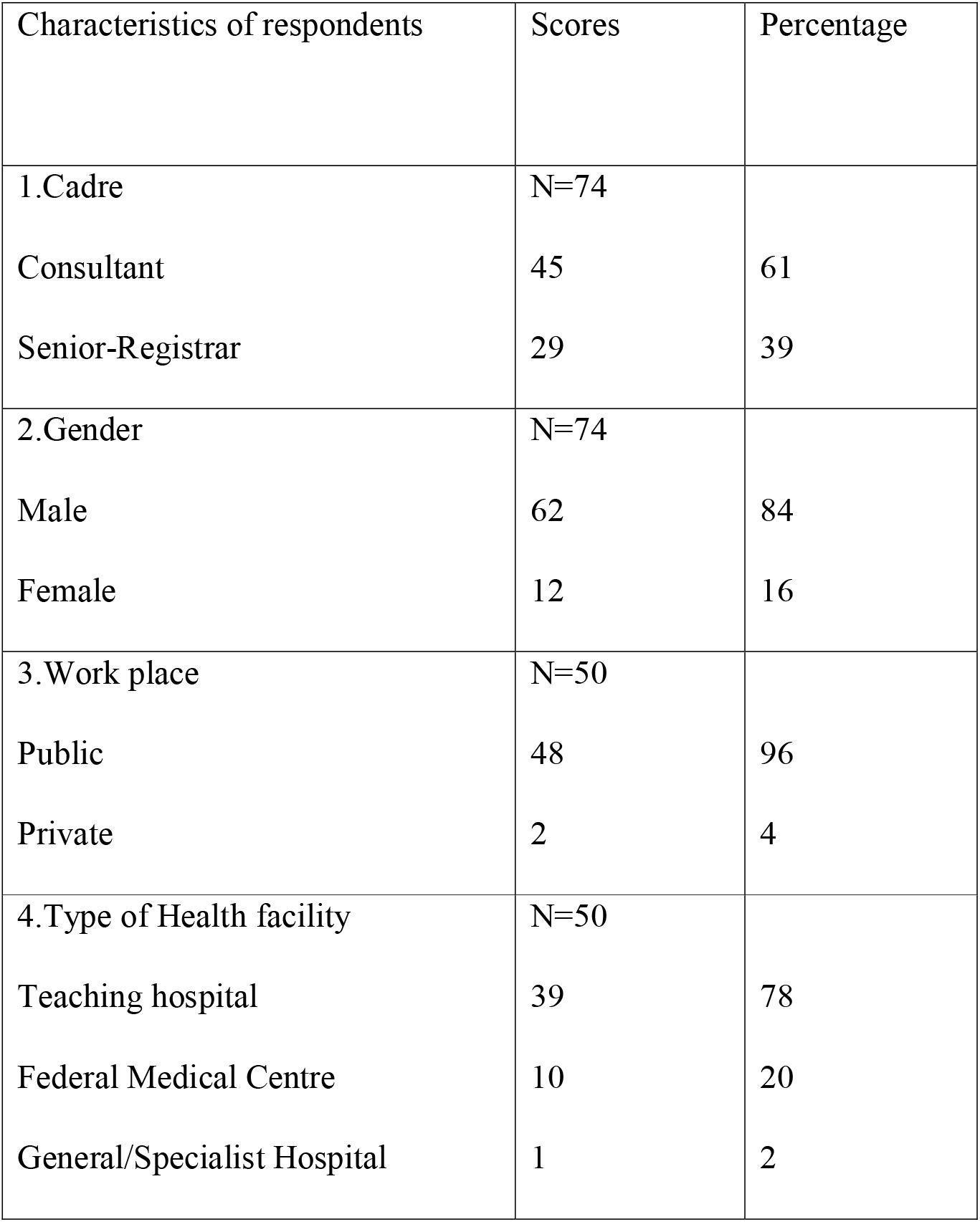
Socio-demographic characteristics of respondents.

### Impact on Surgeries

Elective surgeries had been suspended in 46(92%) centres at the time of this survey. There was a steady decline in the average number of elective surgeries done over 5 months between November 2019 with 993(25% of 5 month total) and March 2020 with 420(10% of total) cases. Similar trend was observed with emergency surgeries which reduced from 822(25% of 5 month total) in November 2019 to 485(15% of total) in March 2020. Comparatively, there were more elective than emergency surgeries per month until March (Figure 1).

**Figure 1:**
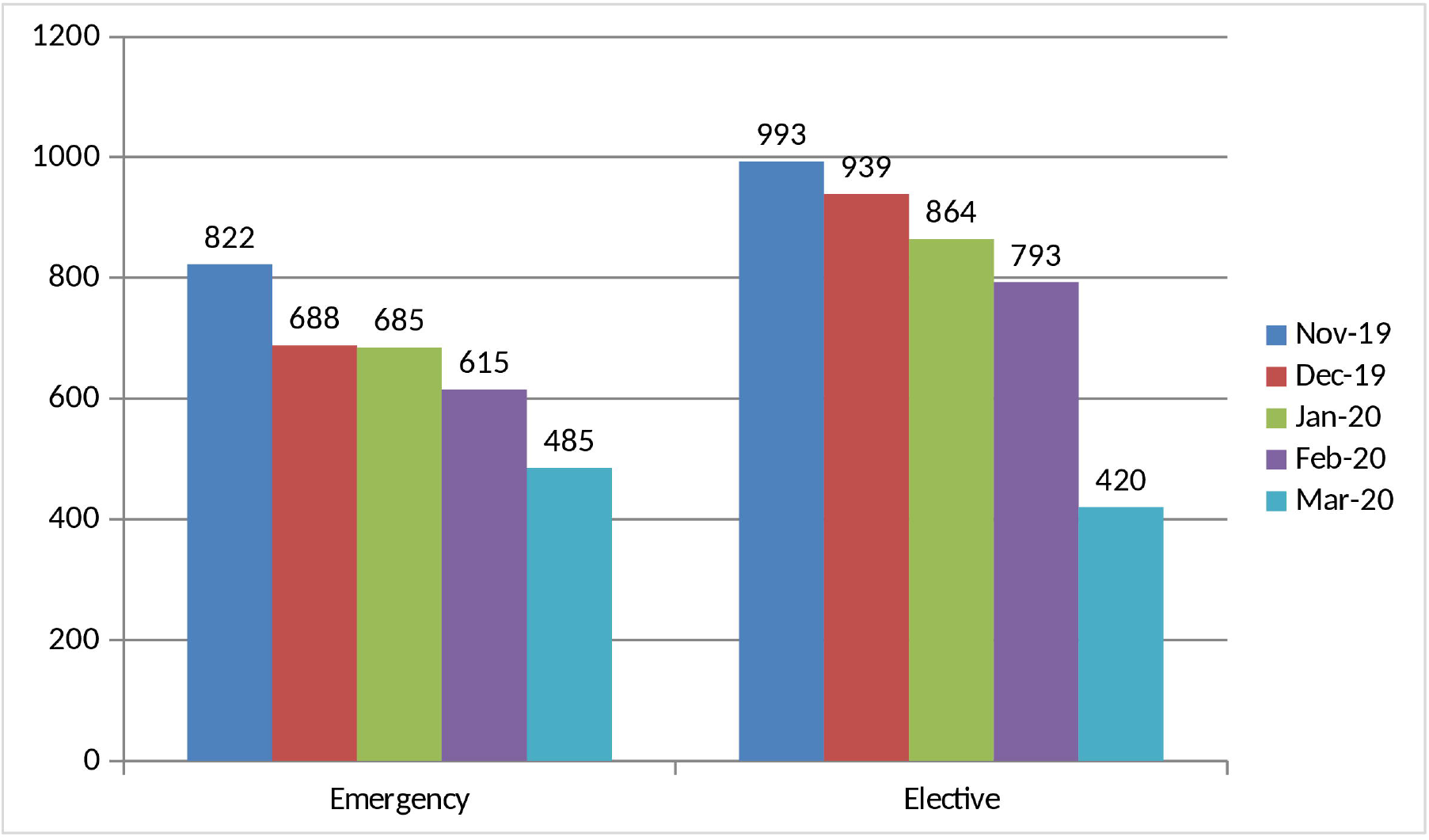
Cluster bar chart of the mean number of surgeries over 5 months.

Twenty (40%) centres suspended their elective surgeries less than 2 weeks prior to the survey in April, 26(52%) centres stopped a month earlier and 4(8%) had suspended their elective list for over a month.

### Adverse clinical outcomes

Eleven (22%) centres reported at least one of the elective cases suspended due to COVID-19 pandemic presenting as emergency in March. There were 13 of such patients accounting for an estimated 3% of the total emergency surgeries for the month. They included inguinoscrotal hernias (10) with obstruction, sub-acute appendicitis (2) and previously decompressing anovestibular fistula with intestinal obstruction (1).

### Changes in Management Modality

Nine (18%) centres have newly adopted non-operative modalities for managing selected surgical conditions in response to the pandemic. One (2%) centre adopted non-operative reduction of intussusception while 5(10%) centres adopted management of uncomplicated acute appendicitis with antibiotics and 3(6%) took a more conservative approach to management of trauma. Three (6%) centres replaced laparoscopic appendectomy with open surgery.

Protocol for the management of urgent cases such as cancers, symptomatic hernias in the early period of COVID-19 was to continue to immediately operate in 31(62%) centres, delayed intervention in 12(24%), masterly inactivity in 2(4%) and follow up in 5(10%).

### Impact on Surgeons

Paediatric surgeons’ perception of safety to operate during the pandemic and their willingness to operate on COVID-19 positive patients are shown in figure 2. Perception of safety to operate rated low in half of respondents. No member of the surgical teams had tested positive for COVID-19.

**Figure 2:**
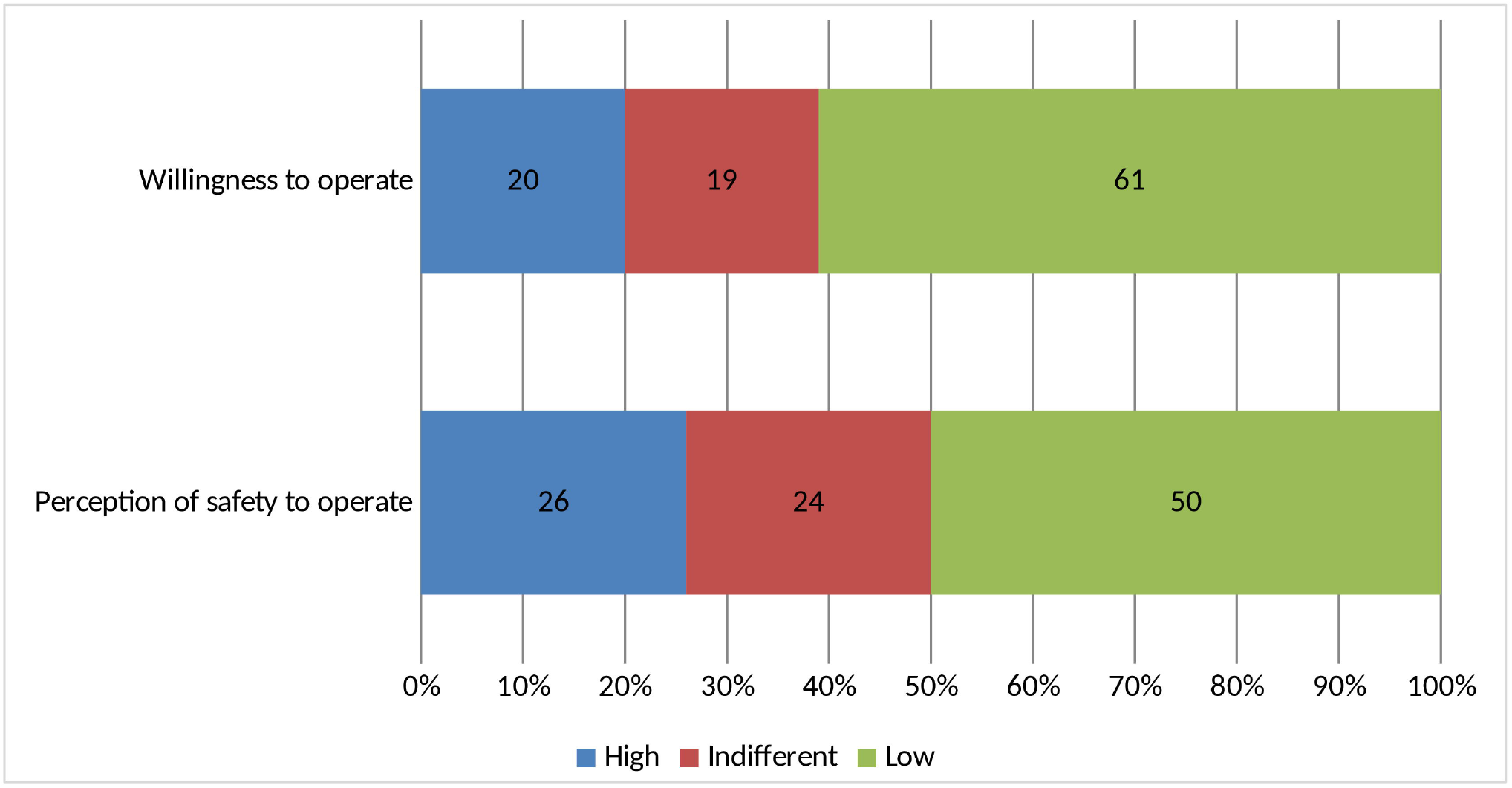
Perception of safety of paediatric surgeons and willingness to operate.

Fifty-seven (77%) agreed to a need for paediatric surgeons to have additional training in management of surgical patients during epidemics, 6(8%) were neutral, while 11(15%) disagreed. Those willing to attend such training were 47(64%), 15(20%) were neutral and 12(16%) were unwilling.

### Impact on Institutions, Supplies and Outpatient Clinics

Forty-two (84%) centres had designated isolation wards but only 2(4%) had COVID-19 positive children on admission and none had managed COVID-19 positive children with surgical condition in their facility at the time of this survey. Majority of centres had low perception of adequacy of PPE for theatre staff both at the time of survey and at 3 months afterwards as depicted in Figure 3. Ratings of how institutions are coping with the COVID-19 pandemic was low in 33(66%), intermediate in 14(28%) and high in 3(6%) centres. Forty (80%) centres do not offer hospital powered telemedicine services for patients follow up despite lockdown on outpatient clinics.

**Figure 3:**
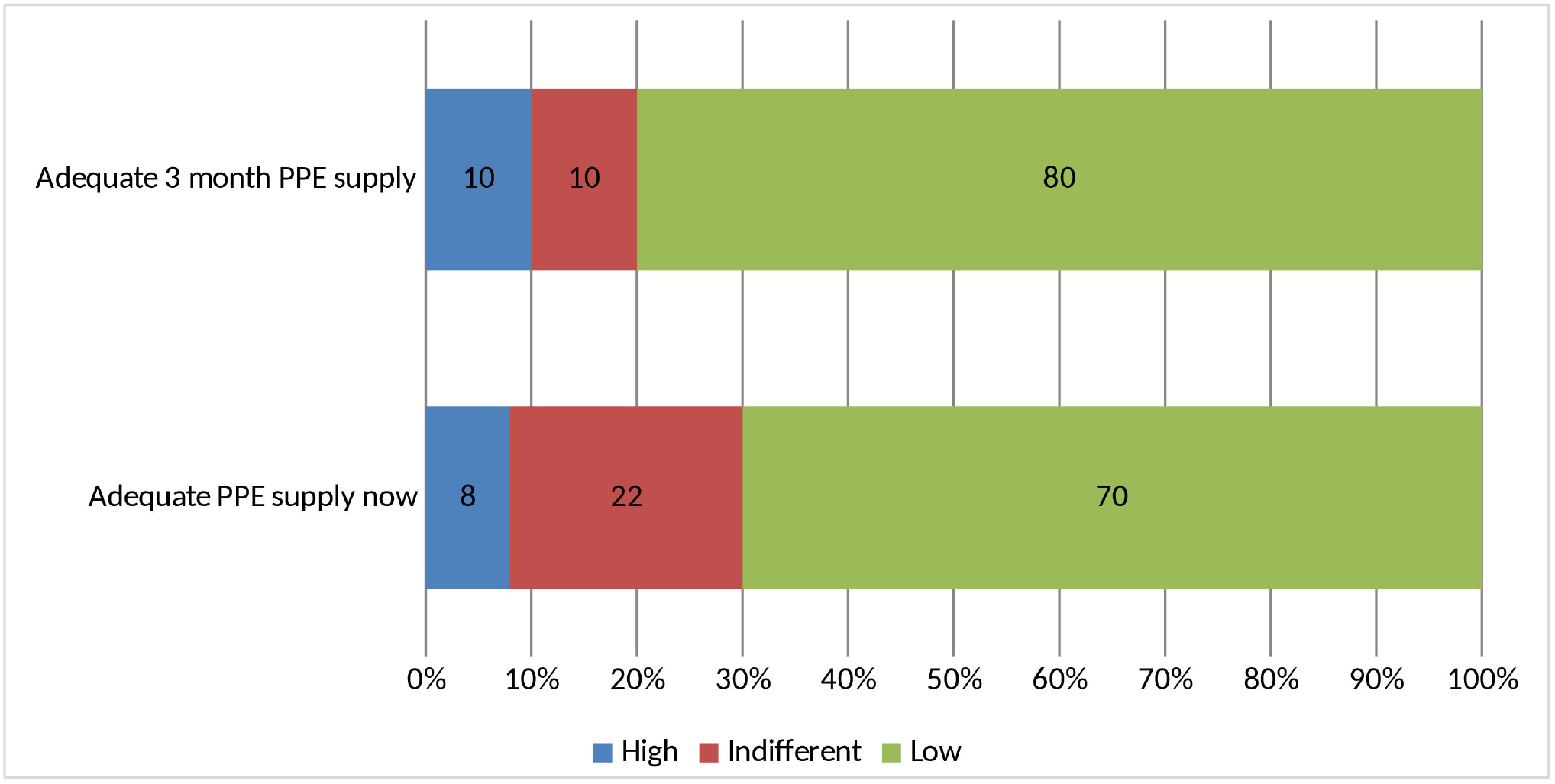
Perception of adequacy of PPE for theatre staff now and in 3 months.

### Impact on Academic Training Programs

Twenty-nine (58%) centres had suspended academic training during the pandemic, 13(26%) engaged “WhatsApp” chat rooms, while 3(6%) made use of Video-conferencing and 5(10%) still carried out their academic training through physical meetings but with social distancing.

## DISCUSSION

Pandemics usually run ravaging course with unpredictable health, social and economic disruptions (4). The impact can be difficult to assess and is an area of active research. While the direct health impact of pandemics can be catastrophic, the indirect impact driven by depletion of resources and reduced assess to routine care can lead to further increase in morbidity and mortality (4).

COVID-19 pandemic is rapidly evolving with unprecedented impact on global health systems. China, and later the United States, Italy and other European countries became hotspots for the virus after reporting their first cases in December 2019 and January 2020 respectively (2,5,6). Travellers from these regions brought in the disease to Africa, including Nigeria in February 2020 (7,8) with a rapid expansion in the number of cases in sub-Saharan Africa (9). The World Health Organization formally declared COVID-19 outbreak a pandemic on 11th March with 634 813 total confirmed cases as at 29th March, 2020 (10,11). This has sparked various adaptations in healthcare responses and management, with unpredictable outcomes heightened by depletion of resources. For example, the only paediatric surgery care facility in Liberia run by Médecins Sans Frontières (MSF) has been temporarily suspended due to travel restrictions (12).

Children are more susceptible to viral respiratory diseases but ironically, statistics on COVID-19 have shown low incidence in this age group. An analysis of 72 314 cases of COVID-19 from the Chinese Centre for Disease Control and Prevention showed a low incidence in children with those younger than 10 years accounting for only 1% of cases (13). A recent observational cohort study of 36 children with COVID-19 found that all the patients had mild (47%) or moderate (53%) type of COVID-19 with large proportion (28%) being asymptomatic (14). This clinical pattern of COVID-19 in the paediatric population could make children important facilitators of viral transmission, and may thus place providers of health care in them at increased risk of infection (14–16).

Our survey showed that majority of the paediatric surgeons have stopped operating on all elective conditions in both public and private tertiary health institutions to minimise contact with potential carriers of the virus and conserve resources. This is consistent with the American College of Surgeons COVID 19: Elective Case Triage Guidelines for Surgical Care which recommended that surgery should be performed only if delaying the procedure is likely to prolong hospital stay, increase the likelihood of later hospital admission or cause harm to the patient (17). A recent article recognises the higher frequency of highly symptomatic patients on the elective operation list in LMICs compared to HICs but still advocates that truly elective operations should be postponed to preserve PPE, staff and facility capacity as important resources during a surge response (18).

The ACS advocates that “children who have failed attempts at medical management of a surgical condition should be considered for surgery” (17). Our study revealed an increased uptake of non-operative management of some surgical conditions such as intussusception, uncomplicated appendicitis and some cases of trauma. This modality of care was probably adopted to reduce exposure to surgery during the pandemic. Outcome of these modifications in management protocol may be subject to future research.

Some suspended elective cases had presented as emergencies. They included incarcerated inguinoscrotal hernias, sub-acute appendicitis and previously decompressing anovestibular fistula that developed partial obstruction. This is an indirect impact of the pandemic due to reduced assess to routine care in these patients. Official tele-medicine platforms for follow up care of patients may aid early detection of complications or other needs for hospital visits while elective surgeries remain suspended, outpatient clinics locked down and patients are being given long appointments. Few centers in our survey have an official tele-medicine platform for follow up care of patients especially during this period of covid-19 pandemic. The ACS recommends that tele-medicine and tele-consult services should be used for patient and physician interaction when available (19).

In this report, all centres continued to operate on emergencies and there was consistent monthly average number of surgeries from November 2019 to February 2020 but a sharp decline in March 2020. This corresponded with the period of social and economic disruptions which followed the first confirmed case of COVID-19 in Nigeria reported on 27th February 2020 (8,20).

Majority of centres had designated isolation wards, but only 4% of them had children with the virus and none had managed a COVID-19 positive child with surgical condition. Although local statistics of incidence in children was not available in our literature search, our finding is suggestive of a low incidence of confirmed COVID-19 in children in Nigeria which is consistent with global data (13,21). Despite this low incidence in children, about half of paediatric surgeons in our survey feel unsafe operating on patients during this period and more are unwilling to operate on confirmed COVID-19 patients.

Majority of centres had suspended academic training during this pandemic. Very few made use of Video-conferencing. Poor internet connectivity and high cost of subscription in sub-Sahara Africa may be partly responsible for this poor uptake of video communication (22,23). Online chat rooms are generally accessible and may be explored as viable media alternative.

This research is survey based with attendant limitation of recall. The study however does provide information on early impact of COVID-19 pandemic on paediatric surgery in Nigeria to help in beginning to plan towards restarting services and handling future unprecedented situations.

## CONCLUSION

The COVID-19 pandemic has resulted in cessation of elective surgeries and a sharp decline in the number of emergency surgeries performed on children in Nigeria. Significant number of pediatric surgeons do not feel safe operating on patients and are mostly unwilling to operate on COVID-19 positive patients in the initial stages of the pandemic. Measures to improve their safety and electronic communication with patients and professional colleagues during the pandemic may help improve the surgical care of children. A follow up study is planned to identify further impacts of the pandemic on children’s surgical care.

## Data Availability

All necessary data for this research have been included in the manuscript.
Additional data are available upon reasonable request from the authors.

## References

1. Lai CC, Shih TP, Ko WC, et al (2020) Severe acute respiratory syndrome coronavirus 2 (SARS-CoV-2) and coronavirus disease-2019 (COVID-19): The epidemic and the challenges. Int J Antimicrob Agents [Internet]. 55(3):105924. Available from: https://doi.org/10.1016/j.ijantimicag.2020.105924

2. Shereen MA, Khan S, Kazmi A, et al (2020) COVID-19 infection: Origin, transmission, and characteristics of human coronaviruses. J Adv Res [Internet]. 24:91–8. Available from: https://doi.org/10.1016/jjare.2020.03.005

3. U.S. Department of Health and Human Services. Pandemic Influenza Plan. 2017 Update [Internet]

4. Pandemics: Risks, Impacts, and Mitigation - Disease Control Priorities: Improving Health and Reducing Poverty - NCBI Bookshelf [Internet]. Available from: https://www.ncbi.nlm.nih.gov/books/NBK525302/

5. Wang C, Horby PW, Hayden FG, et al (2020) A novel coronavirus outbreak of global health concern. Lancet 395(10223):470–3

6. Chiara Severgnini e Redazione Online. Coronavirus, primi due casi in Italia: sono due turisti cinesi - Corriere.it [Internet]. Available from: https://www.corriere.it/cronache/20_gennaio_30/coronavirus-italia-corona-9d6dc436-4343-11ea-bdc8-faf1f56f19b7.shtml?refresh_ce-cp

7. World Health Organization. COVID-19 cases top 10 000 in Africa _ WHO _ Regional Office for Africa. [Internet]

8. Adepopju P (2020) Nigeria responds to COVID-19; first case detected in sub-Saharan Africa. Nature medicine 26:444–448 [Internet]. Available from: http://www.embase.com/search/results?subaction=viewrecord&from=export&id=L631242957%0Ahttp://dx.doi.org/10.1038/d41591-020-00004-2

9. Martinez-Alvarez M, Jarde A, Usuf E, et al (2020) COVID-19 pandemic in West Africa. Lancet Glob Heal [Internet] 2019(20):2019-20. Available from: http://dx.doi.org/10.1016/S2214-109X(20)30123-6

10. World Health Organization. WHO Director-General’s opening remarks at the media briefing on COVID-19. 11 March 2020 [Internet]. WHO Director General’s speeches. 2020. p. 4. Available from: https://www.who.int/dg/speeches/detail/who-director-general-s-opening-remarks-at-the-media-briefing-on-covid-19-11-march-2020

11. World Health Organization. Mariana N (2020) Coronavirus disease 2019 (COVID-19) situation report 69 [Internet]. 2019(3)

12. COVID-19 Interrupts the Only Pediatric Surgery Care in Liberia _ Doctors Without Borders - USA.

13. Zunyou W, Jennifer M (2020) Characteristics of and Important Lessons From the Coronavirus Disease 2019 (COVID-19) Outbreak in China: Summary of a Report of 72 314 Cases From the Chinese Centre for Disease Control and Prevention 323(13)

14. Qiu H, Wu J, Hong L, et al (2020) Clinical and epidemiological features of 36 children with coronavirus disease 2019 (COVID-19) in Zhejiang, China: an observational cohort study. Lancet Infect Dis [Internet] 2019(20):1–8. Available from: http://dx.doi.org/10.1016/S1473-3099(20)30198-5

15. Kelvin AA, Halperin S (2020) COVID-19 in children J: the link in the transmission chain. Lancet Infect Dis [Internet] 2(20):2019-20. Available from: http://dx.doi.org/10.1016/S1473-3099(20)30236-X

16. Fretheim A (2020) The role of children in the transmission of SARS-CoV-2 (COVID-19) - a rapid review [Barns rolle i spredning av SARS-CoV-19 (Covid-19) - en hurtigoversikt] Rapid review, 2020(3): 7–9. Oslo: Folkehelseinstituttet/ Norwegian Institute of Public Health, 2020

17. American College of Surgeons (2020) COVID 19 J: Elective Case Triage Guidelines for Surgical Care. Am Coll Surg. [Internet] 24(3):2020.

18. Weiser TG, Ademuyiwa AO, Capo-chichi N et al (2020) COVID-19 preparedness within the surgical, obstetric and anesthetic ecosystem in Sub Saharan Africa. Ann Surg [Internet]

19. American College of Surgeons. COVID-19 Guidelines for Triage of Pediatric Patients [Internet]. Available from: https://www.facs.org/covid-19/clinical-guidance/elective-case/pediatric-surgery

20. Ehanire O (2020) First case of Coronavirus disease (Covid-19) confirmed in Nigeria. Nigeria Centre for Disease Control. February 28, 2020 [Internet]

21. She J, Liu L, Liu W (2020) COVID - 19 epidemic J: Disease characteristics in children. J Med Virol. 2020(3);1–8. https://doi.org/10.1002/jmv.25807

22. Akue-Kpakpo A (2013) Study on international Internet connectivity in sub-Saharan Africa. International Telecommunication Union Telecommunication Development Bureau Place des Nations CH-1211 Geneva 20 Switzerland www.itu.int [Internet] 2013(3)

23. Concerns over high cost of Internet connection - Punch Newspaper [Internet]. November 4, 2018: 26–7

